# Genomic diversity of respiratory pathogens in clinical versus community surveillance: a comparative analysis of SARS-CoV-2, RSV, and influenza viruses in Washington State

**DOI:** 10.64898/2026.11.11.26367863

**Authors:** Joshua M. Kirsch, Anna Elias-Warren, Lauren Frisbie, Christina Lockwood, Ana A. Weil, Allison Black, Helen Y. Chu, Pavitra Roychoudhury

## Abstract

**Background:** Genomic surveillance of respiratory viruses typically relies on sequences from individuals seeking health care, and whether these sequences represent the viruses circulating in the community is not well-characterized. We aimed to compare the diversity, proportion, and dynamics of influenza, respiratory syncytial virus (RSV), and SARS-CoV-2 between clinical and community samples.

**Methods:** In this retrospective comparative genomic analysis, we used whole-genome sequences from community-based surveillance studies in the Seattle area, and clinical sequences from the Washington State Department of Health. We compared clade proportions and dynamics, sequence-cluster diversity, non-synonymous mutations, and Ct values between clinical and community samples.

**Findings:** We analyzed 538 RSV sequences collected between 2019 and 2021 (278 community, 260 clinical), 1506 influenza sequences (260 community, 1246 clinical) between 2019 and 2020, and 8660 SARS-CoV-2 sequences (5923 community, 2737 clinical) from 2020 to 2022. Clade proportions did not differ between clinical and community samples for RSV or influenza, but differed for SARS-CoV-2: clades 21J (Delta) and 21K (Omicron BA.1) were more frequent in community samples and clade 22B (Omicron BA.5) in clinical samples. SARS-CoV-2 clinical sequence clusters were more diverse than community clusters, whereas RSV and influenza showed no significant difference. Several spike mutations associated with monoclonal antibody resistance were more prevalent in community samples for SARS-CoV-2. C_t_ values were lower in clinical samples for multiple SARS-CoV-2, RSV, and influenza clades.

**Interpretation:** Seasonal respiratory viruses had similar dynamics and diversity in clinical and community samples, whereas pandemic SARS-CoV-2 differed between settings. Genomic surveillance programs should combine clinical and community sampling, particularly during pandemics in order to better capture viral diversity.

**Funding:** This analysis was funded by the Washington State Department of Health Northwest Pathogen Genomics Center of Excellence (WA DOH contract number HED29377-1, federal grant number NU50CK000630 to University of Washington). The underlying studies were funded by Gates Ventures.

**RESEARCH IN CONTEXT:** *Evidence before this study:* Genomic surveillance of respiratory viruses expanded substantially during the COVID-19 pandemic and has since been extended to respiratory syncytial virus (RSV) and influenza, but most sequences come from people who sought medical care. We searched PubMed for studies published in English up to September 2026 using combinations of terms “genomic surveillance”, “community surveillance”, “respiratory syncytial virus”, “influenza”, “SARS-CoV-2”, “whole genome sequencing”, and included studies that sequenced respiratory viruses from defined clinical or community populations. Previous work has sampled hospitalized patients, sequenced random positive specimens, or sequenced wastewater, and has shown that such data can track variant emergence, clade replacement, and nosocomial transmission. However, few studies compared clinical and community sampling for the same virus in the same population and period, so it remains unclear whether clinically derived sequences represent the viruses circulating in the community.

*Added value of this study:* To our knowledge, this is the first study to compare viral diversity between clinical and community sampling for three respiratory viruses in the same region and overlapping time periods. Using more than 10,000 whole genomes from community-based surveillance studies and state clinical surveillance in Washington State, we found that clade proportions and dynamics were similar between sources for RSV and influenza, but differed for SARS-CoV-2. At finer genetic resolution, clinical SARS-CoV-2 sequences formed more diverse clusters than community sequences, and several spike mutations associated with monoclonal antibody resistance were more prevalent in community samples.

*Implications of all the available evidence:* For seasonal respiratory viruses, sampling people who seek care appears to capture much of the diversity circulating in the community, which may allow surveillance programs with limited resources to rely on clinical specimens. In contrast with seasonal circulation in a population with some pre-existing immunity, during pandemic-level transmission, when very large numbers of people in a largely susceptible population are infected over a short period, clinical and community samples are not interchangeable, and programs that sequence only clinical specimens risk missing variants circulating in the community, including mutations relevant to antiviral and monoclonal antibody resistance. These differences may partly reflect the much larger number of SARS-CoV-2 genomes available, as well as heterogeneous mixing and co-circulation of multiple variants during the pandemic. Surveillance programs should therefore combine clinical and community sampling, and future studies should test whether these patterns hold in other regions, seasons, and pathogens.

## INTRODUCTION

Viral whole genome sequencing (WGS) has transformed infectious disease surveillance and outbreak response over the past decade ^1,2^. The ability to rapidly characterize pathogen genetic diversity provides critical insights for public health decision-making, such as tracking transmission chains during outbreaks, identifying emerging variants and monitoring antimicrobial resistance ^3^. Genomic data now routinely complement traditional epidemiological approaches, enabling higher-resolution understanding of disease dynamics than was previously possible with phenotypic or partial gene sequencing methods alone ^4^.

The COVID-19 pandemic catalyzed an unprecedented expansion of pathogen genomic surveillance infrastructure globally ^5^. In the United States, initiatives like the CDC’s Baseline and National SARS-CoV-2 Strain Surveillance programs, combined with substantial investments in state and local public health laboratory infrastructure created and expanded sequencing capabilities across the country allowing genomes to be rapidly shared through public repositories like NCBI GenBank ^6^. This infrastructure has enabled extension of routine genomic surveillance to other respiratory pathogens of public health importance, particularly respiratory syncytial virus (RSV) and influenza (flu) virus ^7^. These three pathogens represent the major viral causes of acute respiratory illness and collectively impose substantial morbidity, mortality, and healthcare burden, especially in the elderly and immunocompromised ^8,9^. In 2019, an estimated 33 million children younger than 5 years contracted RSV and over 101,000 children died due to RSV-associated causes ^10^. Influenza causes infections in 5-10% of adults and 20-30% of children ^11^. RSV genomic surveillance has gained urgency following the licensure of RSV vaccines and monoclonal antibody prophylaxis, creating a need to monitor for immune escape variants^12^.

Viral epidemiology requires population-level surveillance of viral strains, and multiple strategies for measuring the presence of different strains at scale have been utilized for respiratory viruses. This includes sampling specific populations, such as hospitalized patients ^13^, random sequencing of positive samples ^14^, and sequencing of wastewater samples ^15^. Genomic epidemiology of respiratory viruses has identified developments that are crucial for public health responses, such as inter-continental transmission of new pathogenic SARS-CoV-2 variants, nearly complete replacement of RSV clades between seasons, and prediction of emerging variants of influenza virus ^16–18^.

The COVID-19 pandemic was a unique period in which broad, community-level testing was widely available and samples from community members were routinely sent to laboratories for testing and sequencing. As this community testing infrastructure has waned, laboratories generally no longer have access to community samples, and genomic surveillance now relies predominantly on specimens from individuals who present for clinical care. Whether this clinical sampling frame is representative of the viruses circulating in the broader population remains unclear, mainly due to the lack of studies that track viral dynamics in both hospital and community settings ^19^. Past studies have utilized localized wastewater sampling followed by targeted clinical screening to contain SARS-CoV-2 outbreaks ^20^, demonstrating that these data can be highly impactful. Given the high cost of viral genomic surveillance and the reduced funding for the sequencing of respiratory viral isolates post-pandemic, public health agencies need data to decide how to judiciously apply limited resources for the measurement of respiratory virus transmission.

In this study, we analyzed the viral diversity of seasonal RSV and influenza virus strains and pandemic SARS-CoV-2 strains isolated from both community members and from individuals seeking healthcare in the same time frame and geographic location. The overall diversity of SARS-CoV-2 clades differed between sampling modalities but remained similar for RSV and influenza virus, highlighting that pandemic-scale infections in hospitalized populations do not completely reflect the pattern in the community at large. Measurement of the viral diversity over time revealed that intra-clade temporal dynamics also differ between clinical and community infections for SARS-CoV-2. Finally, we found that for SARS-CoV-2, sampling from clinical specimens led to the identification of more diverse strains than sampling from the community, whereas RSV and influenza showed no significant difference. Together, these results suggest that clinical sampling may adequately capture the diversity of seasonal respiratory viruses, but that during pandemic-level transmission, viral populations sampled in hospital settings can diverge from those circulating in the community. Future surveillance programs should prioritize a diverse sampling strategy.

## METHODS

### Study design and setting

We conducted a retrospective comparative genomic analysis of three respiratory pathogens (SARS-CoV-2, RSV, and influenza) using whole genome sequencing data from clinical and community surveillance sources in Washington state. The study period spanned 2019 to 2022, encompassing multiple respiratory virus seasons and capturing periods of varying transmission intensity for each pathogen. Sequences were classified into two surveillance categories based on the context of specimen collection: *clinical surveillance*, comprising specimens collected from individuals seeking medical care at healthcare facilities; and *community surveillance*, comprising specimens collected through population-based sampling programs that did not require participants to be seeking medical attention.

### Data sources

Sequences and metadata were derived from four main sources: the Seattle Flu Study, Seattle Coronavirus Assessment Network, Husky Coronavirus Testing, and publicly available sequences from Genbank.

The **Seattle Flu Study (SFS)** was a multi-arm surveillance study established in 2018 to characterize the genomic epidemiology of respiratory pathogens in the Puget Sound region ^19^. SFS enrolled participants through multiple community-based channels including kiosks at public venues (libraries, transit stations, community centers), home-based self-swab collection, and outreach at schools, workplaces, daycare centers, and homeless shelters. In addition, the clinical arms enrolled individuals seeking care at four regional hospitals and multiple clinics. For this analysis, SFS specimens collected during hospital-based care visits were classified as clinical, whereas specimens collected at outpatient clinics or kiosks set up within these locations (Sea Mar Community Health Centers, UW Hall Health, and Seattle Children’s Hospital outpatient clinic) were classified as community samples. Participants with symptoms associated with acute respiratory illness (ARI) provided nasal swabs specimens and linked information including demographics, symptoms/illness characteristics, and other metadata. Specimens underwent multiplex RT-PCR testing for a panel of respiratory pathogens, with positive samples reflexed to whole genome sequencing for influenza, RSV, and SARS-CoV-2.

The **Seattle Coronavirus Assessment Network (SCAN)** was launched in March 2020 as an extension of SFS infrastructure to enable population-based SARS-CoV-2 surveillance ^21^. SCAN utilized home-based self-collection of nasal swabs, with test kits distributed through mail, drive-through sites, and community partner locations. The program was designed to capture SARS-CoV-2 infections across the community regardless of symptoms, testing access, or healthcare utilization, with enrollment open to all King County residents during the study period. SCAN contributed RSV and influenza sequences from specimens tested on multiplex respiratory panels.

The **Husky Coronavirus Testing (HCT)** program ^22^ provided SARS-CoV-2 screening to students, staff, and faculty at the University of Washington from September 2020 to July 2023. Participants completed daily attestations via email or text message, and those reporting symptoms, recent travel, or exposure to a known SARS-CoV-2 case were offered SARS-CoV-2 testing. Participants could also request testing for any reason in the absence of exposure or symptoms. Participants self-collected nasal swabs at campus testing sites or returned them via campus drop-boxes or courier. Samples were tested by PCR and SARS-CoV-2-positive specimens with Ct values under 30 underwent sequencing using a whole genome amplicon sequencing approach (Illumina COVIDSeq) as described previously ^22^. HCT contributed SARS-CoV-2 sequences to the community surveillance arm of this study.

Together, these studies sampled infections across a spectrum of severity, including asymptomatic and mildly symptomatic cases that would not typically present to healthcare settings. While the programs were concentrated in the Seattle metropolitan area, this geographic focus provides comparability with clinical surveillance data, which was also predominantly from this region.

### Clinical Surveillance

Clinical surveillance sequences for SARS-CoV-2 only were obtained from the Washington State Department of Health (WA DOH) genomic surveillance system, which aggregates sequences from diagnostic testing performed at clinical laboratories throughout the state^14^. The primary sources of clinical sequences included hospital-based diagnostic laboratories, commercial reference laboratories (e.g., LabCorp, Quest Diagnostics), and the University of Washington Clinical Virology Laboratory, which serves as a major reference laboratory for healthcare facilities throughout the Pacific Northwest.

Although the WA DOH surveillance system receives specimens from patients presenting to a range of healthcare settings, including emergency departments, inpatient hospital units, outpatient clinics, and urgent care facilities, the sequences provided by WA DOH for this analysis were specifically from individuals hospitalized as a result of SARS-CoV-2 infection, rather than from all individuals presenting for care. Testing was performed for diagnostic purposes as part of routine clinical care, and specimens were subsequently sequenced through WA DOH’s genomic surveillance program or sequencing efforts at submitting laboratories. The selection of specimens for sequencing varied across the study period and across laboratories, and ranged from sequencing of all positive specimens, random sampling of positives, to targeted sequencing of specimens from hospitalized patients or suspected vaccine breakthrough cases.

Classification of sequences as clinical was performed using the *Washington Disease Reporting System (WDRS)*, WA DOH’s integrated disease surveillance platform. Sequences were classified as clinical if they were linked to a diagnostic test ordered in the context of a healthcare encounter, as documented in WDRS. No additional metadata on these samples was available.

### Study Period and Temporal Considerations

Temporal matching between surveillance sources is critical for valid comparison, as pathogen diversity changes rapidly over time. The study period was selected to maximize temporal overlap between community and clinical surveillance data sources while capturing sufficient genomic diversity for meaningful comparison. For time-stratified analyses, we selected periods with at least 3 months of continuous detection in both surveillance sources. For SARS-CoV-2, the study period of 2020 to 2022 encompasses multiple variant waves, including the emergence and spread of Delta (21J) and the Omicron sub-lineages BA.1 (21K) and BA.5 (22B). The HCT program operated from September 2020 to September 2022, defining the window for community surveillance data availability. For RSV and influenza, the study period of 2019 to 2021 encompasses two respiratory seasons (2019/2020 and 2020/2021), with influenza sequences derived from the 2019/2020 season only, with SFS/SCAN enrollment occurring throughout this period.

### Sequence data and phylogenetic analysis

Specimens from community studies (SFS, SCAN, HCT) were processed and sequenced using standardized protocols within each program (see above). Generally, respiratory specimens were tested by multiplex RT-PCR, and pathogen-positive specimens with cycle threshold (Ct) values below thresholds defined for the study or surveillance program were selected for whole genome sequencing. Sequencing and genome assembly approaches depended on the study, time-period and submitting laboratory, and assembled genomes were deposited to NCBI Genbank. Accessions for sequences used in this analysis are listed in Table S1.

Downloaded sequences were assigned lineages/clades using Nextclade v3.20.0 ^23^ using the “sars-cov-2/wuhan-hu-1/orfs” dataset for SARS-CoV-2, “rsv/a/EPI_ISL_412866” and “rsv/b/EPI_ISL_1653999” datasets for RSV-A and RSV-B respectively, and “flu/h1n1pdm/ha/MW626062”, “flu/h3n2/ha/EPI1857216”, “flu/b/ha/KX058884” for influenza H1N1, H3N2, and B respectively. Genomes with a quality score of less than 100 (corresponding to ‘good’ and ‘mediocre’ quality classes in Nextclade) were included in the final dataset.

Phylogenetic trees were constructed using the Augur pipeline with default settings ^24^. Trees were pruned with treeshrink v1.4.0 ^25^ and visualized using ggtree ^26^. SNP distances were calculated using snp-dists v1.2.0.

### Statistical analysis and data visualization

All statistical analyses were performed in R v4.6.1 and figures were generated using ggplot2 v4.0.3. Differences in the proportional abundance of viral clades between community and clinical samples were assessed using a χ² test of independence, with Cramér’s V reported as a measure of effect size. Differences in clade dynamics over time between the two sampling sources were assessed using a Kruskal-Wallis test, and differences in Ct values between community and clinical samples were assessed using a Wilcoxon rank-sum test. All p-values were adjusted for multiple comparisons using the false discovery rate method. Significance was defined as q < 0.05. Genomic diversity was compared between community and clinical samples using a rarefaction analysis of sequence clusters, defined as genomes within fewer than 5 SNPs of one another [#27 Gregori]. Each population was sampled without replacement 100 times at sequential depths beginning at 10 sequences; the maximum depth sampled equaled the lowest number of sequences per population for each virus. Code used to generate the figures and analyses are available at: https://github.com/uwvirology-ngs/clinical-vs-community-seqs

### Ethical considerations

The Seattle Flu Study and SCAN were approved by the University of Washington Institutional Review Board (IRB) under protocols STUDY00006181 and STUDY00010432, respectively. The Husky Coronavirus Testing program was approved under STUDY00011148. Participants provided informed consent for specimen collection, testing, sequencing, and data sharing.

### Role of the funding source

The funder was not involved in the analysis or in the preparation of the manuscript.

## RESULTS

### An aggregate dataset of clinical and community samples across multiple studies and data sources was assembled for this analysis

The Seattle Flu Study (SFS), Seattle Coronavirus Assessment Network (SCAN), and Husky Coronavirus Testing (HCT) studies were conducted from 2018-2022. In these community-based surveillance studies, we identified and sequenced thousands of respiratory viruses before, during, and after the COVID-19 pandemic. For each specimen, metadata recorded whether the virus was collected from a community participant or from a patient presenting to clinical care. Community sampling occurred through mail-in swab kits, in-person kiosks across the Seattle metropolitan area, and enrollment at schools, workplaces, childcare centers, and shelters (Figure S1). Additionally, the Washington State Department of Health (DOH) provided 1002 SARS-CoV-2 sequences known to be from individuals hospitalized due to SARS-CoV-2 infection (see methods).

For this analysis, we assembled an aggregate dataset containing sequences from these four sources over selected timeframes where there was adequate representation of both community and clinical samples and circulation of the viruses of interest. All sequences were downloaded from NCBI Genbank (Table S1). Our initial dataset included 538 RSV sequences (278 community and 260 clinical sequences, Figure 1A), 1506 flu sequences (260 community and 1246 clinical sequences, Figure 1C) and 8660 SARS-CoV-2 sequences (5923 community and 2737 clinical sequences, Figure 1E. All RSV sequences (100%) and 8624 SARS-CoV-2 (99.6%) sequences passed our filtering criteria (see Methods). RSV sequences were collected during two seasonal spreading events in the winter months of 2019/2020 and 2020/2021 (Figure 1B) and the maximum number of samples collected in a single day was 14 specimens. Flu sequences were collected over a single transmission season in 2019/2020 and the maximum number of samples collected in a single day was 37 (Figure 1D). Conversely, the SARS-CoV-2 isolates were collected during the 2020-2022 SARS-CoV-2 pandemic and the maximum number of samples collected in a single day reached 182 specimens (Figure 1F).

**Figure 1.**
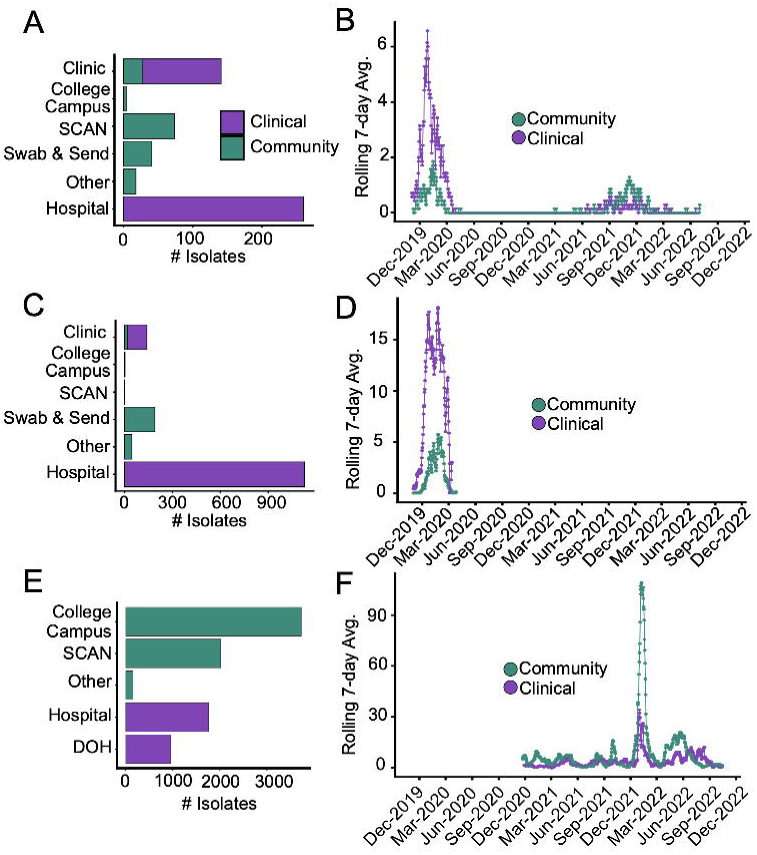
Longitudinal sampling of RSV (A,B), flu (C,D), and SARS-CoV-2 (E,F) from community and clinical sources showing sampling sites (A,C,E) and rolling 7-day mean sampling rates (B, D, F) for each pathogen.

### The dynamics and abundance of SARS-CoV-2 clades differed between clinical and community samples, but no difference was detected among RSV or influenza clades

RSV sequences demonstrated seasonal variation with both RSV-A and RSV-B circulating in the 2019/2020 season, while only RSV-B was detected in the 2020/2021 season (Figure 2A-B). Due to the seasonality of RSV, we first analyzed the overall RSV clade composition in each season. We found no significant difference in the proportion of clades between community and clinical samples (Figure 2A; χ² test with Cramér’s V = 0.027-0.10, all p_adj_ > 0.05). A similar analysis for flu clades also showed no significant differences in clade proportions between community and clinical samples (Figure 3A; χ² test with Cramér’s V = 0.023 – 0.067, p_adj_ = 0.029 - 0.36). These results demonstrate that, at the level of viral clades, RSV and flu viral diversity in hospitalized patients frequently resembles that in community members. In contrast, the proportion of viral clades in SARS-CoV-2 isolates differed significantly between community and clinical samples (Figure 4A; χ² test with Cramér’s V = 0.0069 - 0.22, p_adj_ = 1.25 x 10^-96^ – 0.52). Clades 21J (Delta) and 21K (Omicron BA.1) were isolated more frequently in community samples compared to clinical samples, while clade 22B (Omicron BA.5) was more abundant in clinical samples.

**Figure 2.**
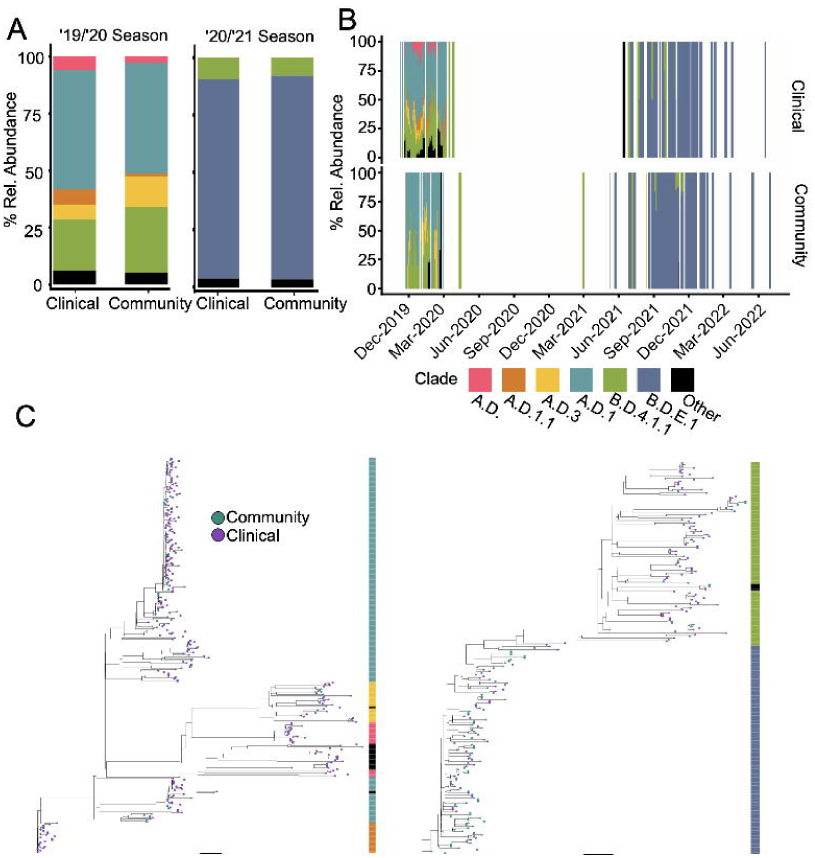
Abundance and dynamics of RSV clades in community and clinical samples. (A) Proportion of viral clades in the 2019/2020 and 2020/2021 season (Chi-squared, * indicates p < 0.05 & Cramér’s V > 0.09). (B) Dynamics of viral clades in each sampling type (Kolmogorov-Smirnov, * indicates p < 0.05 & KS statistic > 0.3). (C) Maximum likelihood trees of RSV-A (left) and RSV-B (right). Only clades with 15 or more sequences are shown.

**Figure 3.**
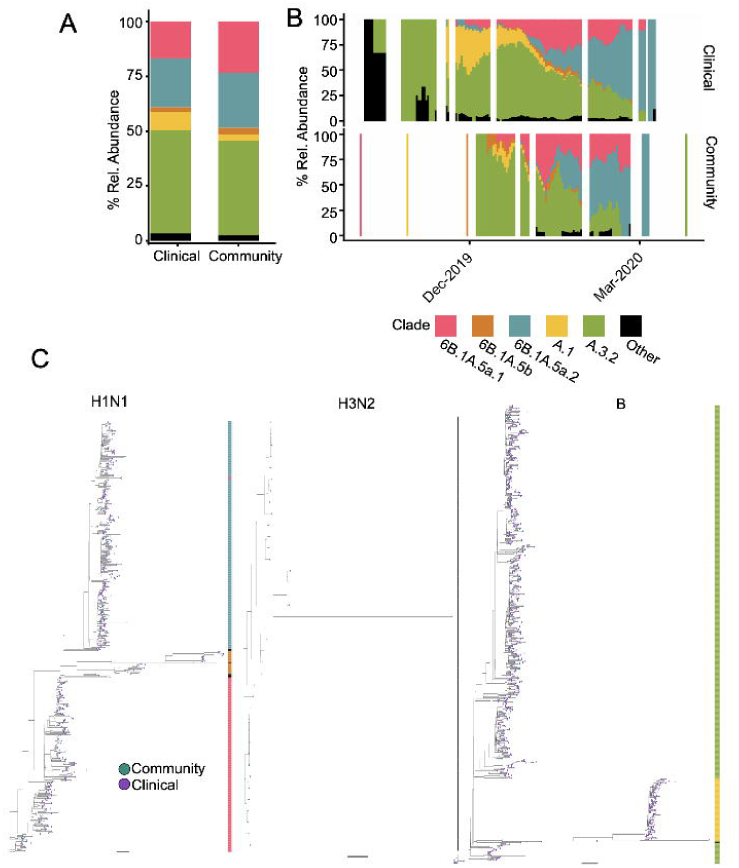
Abundance and dynamics of flu clades in community and clinical samples. (A) Proportion of viral clades (Chi-squared, * indicates p < 0.05 & Cramér’s V > 0.09). (B) Dynamics of viral clades in each sampling type (Kolmogorov-Smirnov, * indicates p < 0.05 & KS statistic > 0.3). (C) Maximum likelihood trees of H1N1 (left), H3N2 (middle), and B (right). Only clades with 15 or more sequences are shown.

**Figure 4.**
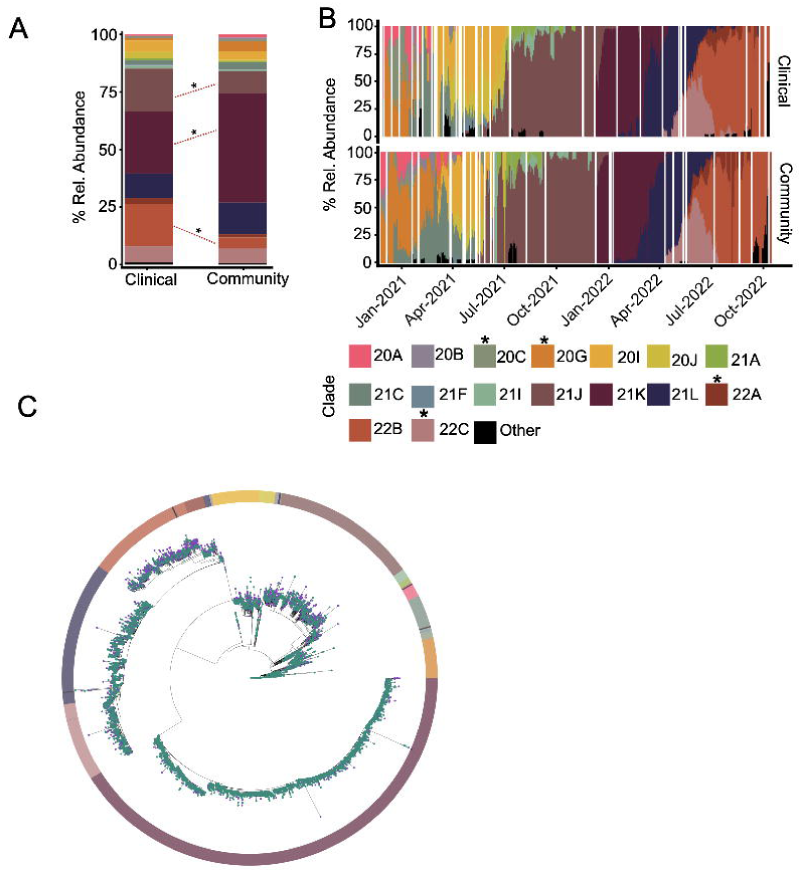
Abundance and dynamics of SARS-CoV-2 clades in community and clinical samples. (A) Proportion of viral clades in the 2019/2020 and 2020/2021 season (Chi-squared, * indicates p < 0.05 & Cramér’s V > 0.09). (B) Dynamics of viral clades in each sampling type (Kolmogorov-Smirnov, * indicates p < 0.05 & KS statistic > 0.3). (C) Maximum likelihood trees of SARS-CoV-2. Only clades with 15 or more sequences are shown.

Next, we compared the temporal dynamics of each clade between clinical and community specimens. For RSV, the dynamics of isolates in the A.D.3 clade were significantly different in clinical and community members (Figure 2B; Kruskal-Wallis test, test statistic = 0.097-0.55 and all p_adj_ > 0.05). No significant differences in dynamics were observed within flu clades (Figure 3B; Kruskal-Wallis test, test statistic = 0.15 – 0.23 and p_adj_ = 0.0147 – 0.22). Comparisons of within-clade dynamics in SARS-CoV-2 sequences demonstrated significant differences within clades 20C (multiple B.1 lineages), 20G (B.1.2), and 22C (Omicron BA.2.12.1) (Figure 4B; Kruskal-Wallis test, test statistic = 0.11 - 0.65 and p_adj_ = 1.25 x 10^-14^ - 0.54).

### Clinical and community sequences intermix phylogenetically, but clinical SARS-CoV-2 clusters are more diverse

We next examined diversity at finer genetic scales by examining clustering of community and clinical sequences on phylogenetic trees. In maximum likelihood phylogenies of RSV-A and RSV-B (Figure 2C), and similarly in flu phylogenies for H1N1, H3N2, and B (Figure 3C), and SARS-CoV-2 (Figure 4C), we observed high level of intermixing between community and clinical isolates suggesting that these sequences derived from common ancestors.

Next, we hypothesized that clinical infections presented a bottleneck to diversity because only some infections progress to a severity warranting hospitalization and the viruses causing these infections are selected from a bigger, more diverse pool of viruses in the community. To test this, we compared the diversity of sequence clusters (genomes within <5 SNPs of one another) between clinical and community sequences through a rarefaction analysis, which accounts for differences in sample sizes through repeated sampling ^27^. Rarefaction curves of RSV and FLU clusters revealed no strong difference between sources (Figure 5A-B). However, SARS-CoV-2 clinical sequence clusters were more diverse than community sequences (Figure 5C). This suggests that sampling hospitalized patients reveals greater genetic diversity than sampling pandemic viruses in the community, possibly because participants in community screening were more homogeneous in their exposures, and thus sampled fewer transmission chains, than individuals hospitalized for SARS-CoV-2 infection.

**Figure 5.**
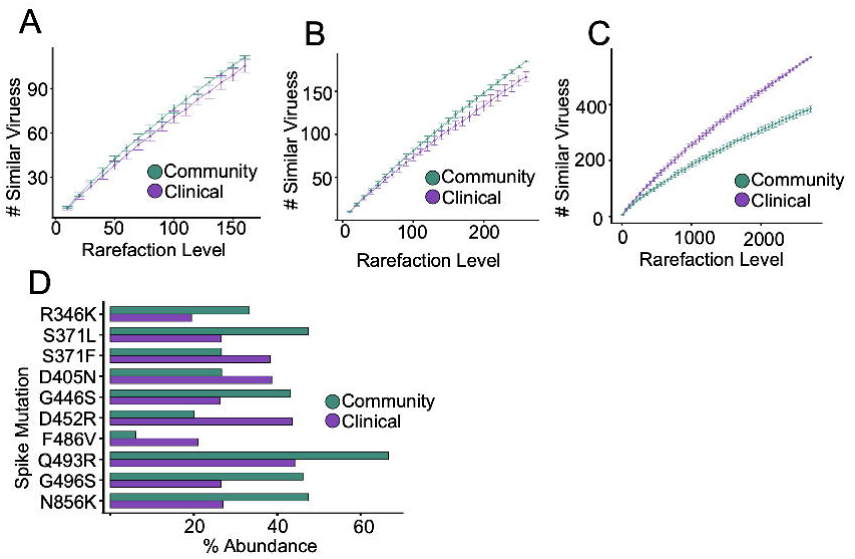
Nucleotide- and amino acid-level diversity of respiratory viruses. Rarefaction curve of genomic clusters (<5 SNPs per cluster) of RSV (A), flu (B), and SARS-CoV-2 (C). (D) Significantly enriched SARS-CoV-2 spike mutations associated with antiviral resistance (* indicates p < 0.05 & Cramér’s V > 0.09).

### Non-synonymous substitutions, including antibody-resistance mutations, differ in frequency between clinical and community samples for RSV and SARS-CoV-2 but not flu

We then compared the abundance of non-synonymous amino acid mutations in flu, RSV, and SARS-CoV-2 between clinical and community populations, as single substitutions can alter drug resistance or pathogenicity. For RSV (Table S2), 37 non-synonymous mutations differed significantly in frequency between the two sources (n = 373 clinical and 165 community sequences; χ² test with Cramér’s V = 0.13–0.32, all p_adj_ < 0.05). All 37 were in RSV-A and all were enriched in clinical samples, concentrated in the attachment glycoprotein G (15 mutations) and the polymerase L (14 mutations); the largest differences were at G:I265L (66.2% of clinical versus 31.5% of community sequences) and L:L1438Q (66.5% versus 32.7%; p_adj_ = 4.7 × 10□¹²). For SARS-CoV-2 (Table S3), 80 mutations differed significantly between sources (n = 2,717 clinical and 5,907 community sequences; Cramér’s V = 0.09–0.24, all p_adj_ ≤ 2.0 × 10□¹□), but enrichment was bidirectional with 51 mutations more frequent in clinical samples and 29 in community samples. The strongest clinical enrichments were S:L452R (43.6% versus 20.1%; Cramér’s V = 0.24, p_adj_ = 4.6 × 10□¹¹¹), S:F486V (21.1% versus 6.2%) and M:D3N (18.4% versus 4.7%), whereas spike changes characteristic of Omicron BA.1, including S:Q493R (44.2% versus 66.6%), S:L212I and S:ins214EPE (both ∼26% versus ∼47%), were more frequent in community samples. The bidirectional pattern for SARS-CoV-2 is consistent with the clade-composition differences described above, whereas the uniformly clinical enrichment in RSV-A was not accompanied by a detectable difference in clade proportions. For flu, no mutations differed significantly in frequency between clinical and community sequences.

We extended this analysis by comparing the enrichment of SARS-CoV-2 spike protein mutations associated with monoclonal antibody resistance (Figure 5D; χ² test with Cramér’s V = 0.11 – 0.24, p = 4.55 x 10^-111^ - 2.22 x 10^-27^). We hypothesized that antiviral resistance mutations would be preferentially enriched in clinical isolates due to the higher usage of antivirals in this population. Surprisingly, certain mutations (R346K, S371L, Q493R, G496S, N856K) were more prevalent in community samples. These results demonstrate that clinically relevant mutations can be widespread in the community.

### Clinical samples have higher viral loads than community samples

Given that clinically attended infections are more severe compared to infections in community members, we hypothesized that the C_t_ value (inversely proportional to number of viral genome copies) in clinical samples would be lower than in community samples. We compared the range of C_t_ values per clade for the subset of sequenced samples where both the C_t_ values and lineage assignments were available for flu (1462, 97%), RSV, (538, 100%), and SARS-CoV-2 (7560, 85.3%). RSV clades A.D.1 and B.D.4.1.1 had significantly lower C_t_ values in clinical samples compared to community samples (Figure 6A, Wilcox ranked-sum test, p = 0.028 - 0.035). The distribution of influenza clades 6B.1A.5a.1, 6B.1A.5a.2, and A.3.2 (Figure 6B, Wilcox ranked-sum test, p = 9.01 x 10^-17^ - 2.03 x 10^-9^) and multiple SARS-CoV-2 clades (Figure 6C, Wilcox ranked-sum test, p_adj_ = 5.51 x 10^-146^ - 6.76 x 10^-3^) showed a similar pattern. No clade had significantly lower Ct values in community samples. These results demonstrate that among sequenced samples, clinically attended respiratory viral infections often have higher viral load.

**Figure 6.**
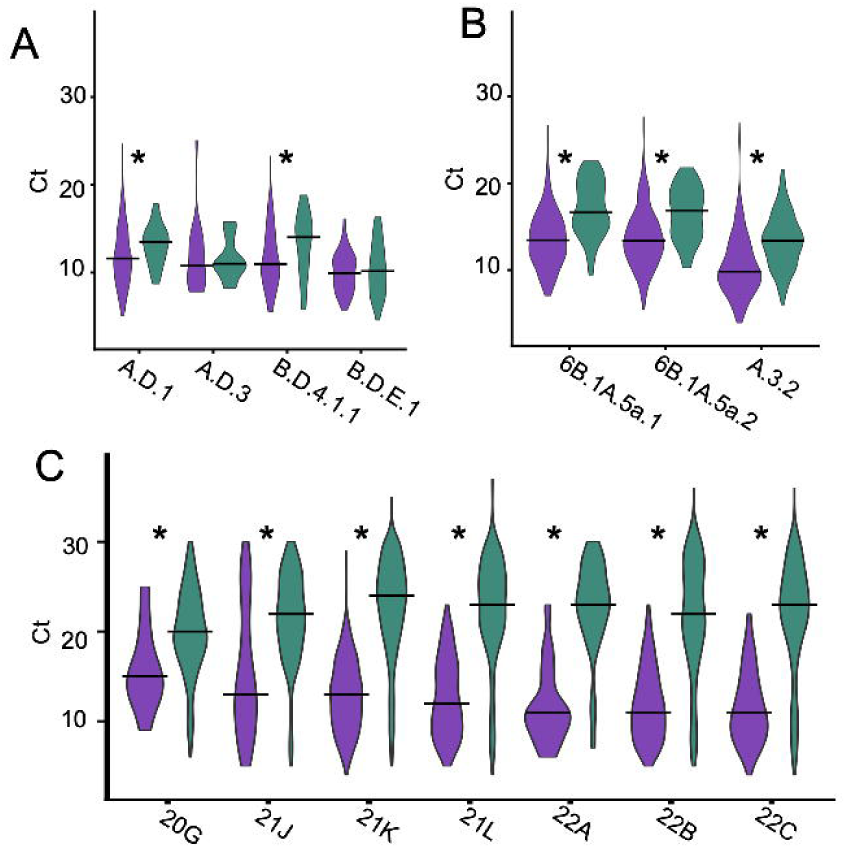
Comparison of Ct values between community and clinical Isolates in (A) RSV, (B) flu, and (C) SARS-CoV-2 clades (Wilcoxon test, * indicates p < 0.05). Only clades with at least 10 sequences from each population were included.

## DISCUSSION

Respiratory virus surveillance often aims to identify new variants which may have increased drug resistance or pathogenicity. Most sequences in public repositories, however, come from specimens collected when people seek medical care, as opposed to community-based programs that sample individuals irrespective of care-seeking. It is unclear how well either source of sampling represents the true diversity of viruses circulating in a population. In this work, we compared multiple levels of viral diversity between samples acquired from community and clinical sampling programs for RSV, flu, and SARS-CoV-2.

We found that high-level viral diversity at the level of clades was significantly different between these two sources for SARS-CoV-2 isolates but not for RSV or flu. This could be due to number of factors. First, the transmission and spread of SARS-CoV-2 during our sampling period was a pandemic-level mass infection of a susceptible population while RSV transmission occurred in a seasonal, periodic manner in a population with some level of pre-existing immunity. RSV transmission also occurred after suppression from the COVID-19 pandemic and before anti-RSV monoclonal antibodies were available. Basic transmission differences such as the number of people being infected and those who are more likely to be severely infected, such as infants with RSV ^28^, may factor into this difference. RSV, influenza, and SARS-CoV-2 are also genetically distinct viruses that replicate through different mechanisms. Another contributing factor to this finding could be the unequal sampling between clinical and community sites, especially for the SARS-CoV-2 isolates ^29^. Oversampling certain groups, for example on college campuses, would yield more closely related viruses, underestimating community diversity and skewing community clade frequencies toward locally circulating strains. Differences in sample size may also increase power to detect clade differences for SARS-CoV-2, and lower-viral-load community infections may be underrepresented among sequenced samples because sequencing success depends on viral load.

We hypothesized that severe infections would result in a bottleneck and therefore expected within-clade diversity to be lower in clinical samples. However, rarefaction analysis of genomic clusters showed the opposite pattern for SARS-CoV-2, with clinical sequences more diverse than community sequences, and no clear difference between sources for RSV or flu. One explanation for this may be that individuals hospitalized for SARS-CoV-2 infection were drawn from a broader cross-section of the population than participants in community testing programs, as noted above, and therefore captured a wider range of transmission chains. Another is that clinical infections are associated with a higher viral load, which likely increases the intrapersonal diversity of viral strains in an individual and a greater composite divergence from the original infecting strain. Nosocomial spread of respiratory viruses is well-documented, and multiple studies have found direct lines of transmission of RSV and SARS-CoV-2 between patients using genomic analyses; however, such transmission would be expected to enrich a few outbreak strains and lower diversity, and WA DOH sequences came from individuals hospitalized because of SARS-CoV-2 infection rather than infected after admission. ^30–32^

The number of mutations between viral clades can be large and accumulation of these mutations can occur slowly, while single substitutions within a viral clade can rapidly lead to drug resistance or increased pathogenicity ^33,34^. We therefore compared the frequency of individual non-synonymous substitutions, including mutations associated with monoclonal antibody resistance, between clinical and community sequences for each of the three viruses. At this resolution the two sources also diverged, but not in the direction expected. RSV showed non-synonymous mutations enriched in clinical sequences despite indistinguishable clade proportions, while SARS-CoV-2 showed differentially abundant mutations with enrichment in both directions, and no flu mutations differed significantly. Several spike substitutions associated with monoclonal antibody resistance were in fact more prevalent in community samples, indicating that clinically relevant mutations circulate widely outside of care settings and may be detected late by clinical sampling alone.

This study has a number of limitations. First, sampling differed markedly across programs in geography, demographics, symptom status, and specimen-selection criteria; although rarefaction addresses unequal depth through repeated sampling at sequential levels of observations, it cannot fully remove these other biases. Second, the comparison rests on three viruses sampled in one region over a limited window, including the atypical pandemic period, which perturbed RSV seasonality and may limit generalizability. Third, assembly and specimen-selection methods varied among the laboratories contributing clinical sequences, which we mitigated with uniform quality filtering but could not fully standardize. Fourth, Ct values were available for only a subset of specimens and are sensitive to sampling timing and swab type. Fifth, the level of sequences available for each virus differed greatly. This can inflate the power of analyses used here and confounds direct comparisons between each virus. Sixth, clade definitions differ in granularity across the three viruses: the Nextclade nomenclature for SARS-CoV-2 partitions diversity into more, finer-scale clades than those currently defined for RSV and influenza, so more within-clade diversity is collapsed for the latter. Finer partitioning increases the opportunity to detect differences in clade composition between sampling sources, which may partly contribute to the larger differences observed for SARS-CoV-2. Our cluster-based rarefaction analysis, which uses a uniform SNP threshold independent of clade nomenclature, partially mitigates this concern.

In conclusion, this study demonstrates key differences in viral dynamics between clinical and community-based sampling and suggests that a combined approach is best for capturing both the breadth and depth of viral diversity. However, when resources are limited, more constrained studies that sample hospitalized patients will likely recapitulate the diversity seen in the community or even surpass it.

## Data Availability

All data produced in the present work are contained in the manuscript and linked to repositories.

https://github.com/uwvirology-ngs/clinical-vs-community-seqs

## CONTRIBUTORS

H.Y.C., A.E.-W., A.B., and P.R. conceptualized the study and obtained funding for the analysis. H.Y.C., A.A.W., A.E.-W., L.F. contributed specimens, sequence data, and/or associated metadata. J.M.K. performed the bioinformatic, phylogenetic and statistical analyses, and prepared the initial figures. J.M.K. and P.R. wrote the original draft and revised figures and text. All authors reviewed and edited the manuscript and approved the final version. J.M.K., A.E.-W., and P.R. directly accessed and verified the underlying data reported in this manuscript. All authors had full access to all the data in the study. P.R. and H.Y.C. had final responsibility for submission.

## DECLARATION OF INTERESTS

H. Y. C. has served on advisory boards for Vir and Roche. P.R. reports contract testing and consulting agreements to UW with Aicuris and Arisan Therapeutics outside of the submitted work. A.A.W. reports research funding from Pfizer, serving on the advisory board at PhagePro, and consulting with General Medicine and serves as a peer reviewer for UpToDate. All other authors report no potential conflicts.

## DATA SHARING

All sequences analyzed in this study are available in NCBI GenBank, with accessions of included sequences provided in Table S1. Analysis code is available at https://github.com/uwvirology-ngs/clinical-vs-community-seqs.

## ACKNOWLEDGMENTS

We thank Chidozie Declan Iwu, Sally Grindstaff, and Jeremy Stone from the University of Washington for assistance with data and troubleshooting. We thank investigators, staff, and participants of the Seattle Flu Study, Seattle Coronavirus Assessment Network, and Husky Coronavirus Testing Program for their contributions to the datasets underlying this analysis. This analysis was funded by the Washington State Department of Health Northwest Pathogen Genomics Center of Excellence (WA DOH contract number HED29377-1, federal grant number NU50CK000630 to University of Washington). AI tools were used only in proof-reading and editing the manuscript for clarity and readability. All edits were reviewed by the authors, and the authors take full responsibility for the content of this publication.

## Notes

### Author Declarations

The University of Washington Institutional Review Board gave ethical approval for this work.

